# Patient and Physician Perspectives on Cardiovascular Risk: A Multicenter Study of Communication Gaps

**DOI:** 10.1101/2024.12.02.24318359

**Authors:** Juan Górriz-Magaña, Ramon Maruri-Sánchez, Ane Elorriaga, Nahikari Salterain-González, Alicia Prieto-Lobato, Raúl Gascueña Rubia, Isabel Monedero Sánchez, Ana Elvira-Laffond, Miguel Lapena Reguero, Amanda Leandro Barros, Cristina Villabona Rivas, Alejandro Gutiérrez Fernández, César Jiménez Méndez, Silvia Prieto-González, María Melendo-Viu, Blanca Alcón Durán, Emilio Blanco López, Clara Bonanad Lozano, Alejandro Durante-López, Anna Carrasquer, Pedro Martínez Losas, Teresa Alvarado Casas, Pedro Pájaro Merino, Victor Juárez Olmos, Javier Lopez-Pais, Michelle M Kittleson, Pablo Díez-Villanueva, Jordi Bañeras-Rius

**Affiliations:** Cardiology Department, Central de la Defensa Gómez-Ulla University Hospital. Madrid, Spain; Cardiology Department, Fundación Jiménez-Díaz University Hospital. Madrid, Spain; Cardiology Department, University Hospital of Basurto. Bilbao, Spain; Cardiology Department, Clínica Universidad de Navarra. Pamplona, Spain; Cardiology Department, Hospital General Universitario de Albacete. Albacete, Spain; Cardiology Department, Severo Ochoa University Hospital. Leganés, Spain; Cardiology Department, University Hospital Fundación de Alcorcón. Alcorcón, Spain; Cardiology Department, University Hospital of Salamanca. Salamanca, Spain; Cardiology Department. Complejo Hospitalario Universitario de A Coruña. A Coruña, Spain; Cardiology Department, Hospital Universitario Príncipe de Asturias. Alcalá de Henares, Spain; Cardiology Department, Hospital Universitario de Navarra. Pamplona, Spain; Cardiology Department, Hospital San Pedro. Logroño, Spain; Cardiology Department, Hospital Universitario Puerta Del Mar. Cádiz, Spain; Cardiology Department, San Agustín University Hospital. Avilés, Spain; Cardiology Department, Alvaro Cunqueiro University Hospital. Vigo, Spain; Cardiology Department, Hospital Vithas Madrid La Milagrosa. Madrid, Spain; Cardiology Department, Hospital General Universitario de Ciudad Real. Ciudad Real, Spain; Cardiology Department, Hospital Clínico Universitario de Valencia. Valencia, Spain; Cardiology Department, Doce de Octubre University Hospital. Madrid, Spain; Cardiology Department, Hospital Universitario Joan XXIII. Tarragona, Spain; Cardiology Department, Infanta Leonor University Hospital. Madrid, Spain; Cardiology Department, La Luz University Hospital. Madrid, Spain; Cardiology Department, Juan Ramón Jiménez University Hospital. Huelva, Spain; Cardiology Department, La Paz University Hospital. Madrid, Spain; Cardiology Department, University Clinical Hospital of Santiago de Compostela. Santiago de Compostela, Spain; Cedars-Sinai Heart Institute; Cardiology Department, Hospital De La Princesa. Madrid, Spain; Cardiology Department, Hospital Universitario Vall D’Hebrón. Barcelona, Spain

**Keywords:** Communication, Risk, Cardiovascular, Information, Procedural, Care

## Abstract

**Background:** Effective risk communication is crucial for managing cardiovascular disease, the leading cause of global mortality. Clear communication between patients and physicians is essential for informed decision-making, yet gaps in understanding often persist.

**Objective:** This study examines the state of risk communication during hospital admissions for cardiovascular events, evaluating both patient and physician perspectives and identifying factors influencing risk perception.

**Methods:** The SEC-HARIPA study is a multicenter, cross-sectional analysis conducted across 28 Spanish centers from October 2022 to March 2023. It included 943 patients admitted to cardiology departments, either urgently or for scheduled procedures. Patients and their physicians completed parallel questionnaires assessing perceptions of cardiovascular risk, procedural complications, and overall communication. Statistical analysis focused on agreement between patient and physician perceptions and the impact of demographic and clinical variables.

**Results:** The mean age of participants was 68.2 years, with ischemic heart disease being the most common cause of admission (41.3%). While 76.9% of patients felt adequately informed about procedural risks, 69.3% of those experiencing adverse events reported a lack of information on potential complications. A significant discrepancy was found between patients’ and physicians’ perceptions of future cardiovascular risk, with patients often underestimating their risk (weighted kappa index of 0.29). Key factors influencing risk perception included prior cardiovascular history, age, gender, and diabetes mellitus.

**Conclusions:** This study reveals significant gaps in risk communication in cardiovascular care, particularly regarding future risks and complications. Tailoring communication strategies to individual patient characteristics could improve understanding and align perceptions with clinical realities, enhancing health outcomes.

## Introduction

Risk communication (RC) is fundamental in health communication practice(Bakhit et al., 2024;Austin & Fischhoff, 2012) particularly in the context of physician-patient relationship and cardiovascular diseaes, which remain the leading cause of global mortality ^3^. This form of communication involves the exchange of information, advice, and opinions regarding risks, encompassing not only the patient’s illness, but also recommended medical proceesdures and prescribed medications^4^.

The primary objective of this communication process is to empower patients with a comprehensive understanding of their health status, facilitating informed decision-making concerning their treatment and management^5^. Consequently, well-informed patients are more inclined to adopt preventive measures and potentially achieve better health outcomes ^1,6^.

However, effective RC is not an inherent skill; rather, it is an acquired ability through knowledge, preparation, training, and practice^7^. It goes beyond merely presenting statistical data and probabilities, emphasizing the implementation of effective communication strategies ^8^.

Numerous factors can contribute to suboptimal RC ^9^ These include healthcare professionals lacking training in communication skills ^10^ as well as time constraints that hinder adequate discussion of risk-related concerns and the ability to foster a trusting environment where patients can freely express their doubts, worries, and expectations^11^. The use of technical jargon instead of plain and understandable language can further impede effective communication^12,13^. Additionally, the absence of open, honest, and non-judgmental dialogue and the transmission of contradictory information from various physicians may also hinder the process^14^. Moreover, when information is conveyed in purely statistical terms without personalized recommendations tailored to each patient’s life, emotional responses such as undue anxiety, stress, or unwarranted optimism can arise^15,16^.

Given these challenges, RC entails more than just presenting information; it also requires ensuring that patients have comprehended and processed the information to make well-informed decisions ^17^. This aspect is critical for achieving appropriate primary and secondary prevention in the context of cardiovascular diseases ^18^.

Despite its paramount importance, there is limited research on how cardiologists effectively communicate risk information to patients. Thus, the present study seeks to assess the current state of RC in the context of hospital admission due to cardiovascular diseases, evaluate patients’ risk perceptions related to physicians’ perception and analyze factors that could affect that relationship.

## Methods

The SEC-HARIPA study is a multicenter, observational, cross-sectional, and descriptive study conducted at 28 Spanish centers between October 2022 and March 2023.

### Study Population

The study comprised consecutive patients aged 18 and above, admitted to the Cardiology department (ward or cardiac intensive care unit) either urgently or as part of a scheduled procedure. Exclusion criteria were limited to individuals with physical or mental conditions that hindered questionnaire completion. The research protocol received approval from the Ethics Committee of the Central University Hospital of the Defense “Gómez Ulla”, as well as from the Ethics Committees of the participating hospitals, and adhered to the Helsinki Protocol.

### Risk Communication Questionnaire

The questionnaire employed in this study was adapted from the “Improving Cardiovascular Risk Communications” (see Supplementary Method 1) toolkit developed by the American College of Cardiology (ACC, © 2017 American College of Cardiology Foundation)^19^ (See Supplementary Method 2). This toolkit provides a comprehensive framework for assessing patients’ perceptions of cardiovascular risk events, associated emotions, and any residual uncertainties they may have. To ensure the questionnaire’s accuracy and cultural relevance, it underwent a meticulous translation process. This included a direct translation from English to Spanish followed by back-translation, all of which was carefully overseen by bilingual physicians. The final version was approved by both our research team and ACC members to maintain the integrity and fidelity of the original content.

To further enhance the questionnaire’s utility, we incorporated additional context-specific questions aimed at deepening our understanding of patients’ responses. These questions covered a range of topics including demographic information, the specific cardiovascular condition leading to admission, risks associated with procedural complications, history of previous cardiovascular admissions and conditions, and various cardiovascular risk factors.

Furthermore, a parallel questionnaire was developed for the treating physicians. This was designed to capture their perspectives on the patient’s admission diagnosis, the likelihood of future cardiovascular events, and whether they believed that patients had been adequately informed about the planned interventions during their hospital stay.

### Questionnaire Administration

The questionnaires were administered to patients during their hospital stay, specifically after their clinical condition had stabilized and within 24 hours prior to discharge. This timing ensured that patients had received comprehensive information about the reasons for their admission and any procedures they had undergone. Before completing the self-administered questionnaire, patients were provided with a clear explanation of the informed consent process to ensure they fully understood their participation. The questionnaire was provided in a paper format, and patients completed it independently without supervision.

On the same day, the treating physicians independently completed their corresponding questionnaires, allowing for a direct comparison between the patients’ perceptions and the physicians’ assessments. This synchronized approach aimed to capture an accurate and contemporaneous reflection of the communication and understanding between patients and their healthcare providers.

### Statistical Analysis

Quantitative variables were reported as mean ± standard deviation, while categorical variables were expressed as percentages. To evaluate the degree of agreement between patients’ and physicians’ perceptions, the kappa index was employed for nominal categorical variables. This index, which ranges from −1 to 1, measures interrater agreement, where 1 indicates perfect agreement, 0 signifies agreement by chance, and −1 denotes perfect disagreement. For ordinal categorical variables, the weighted kappa index was used, which assigns more weight to agreements or disagreements that are closer in rank, thereby providing a more nuanced assessment of correlation when categories have a natural order.

The study aimed to achieve three primary objectives. First, we analyzed patients’ perceptions of their cardiovascular risk, using means and standard deviations for continuous variables and percentages for categorical variables. Second, we evaluated the correlation between patients’ and physicians’ perceptions to understand the alignment—or lack thereof—between the two. Finally, we identified and described key factors that may influence patients’ risk perception, considering both demographic and clinical variables. This comprehensive approach allowed us to assess not only the current state of risk communication but also the underlying factors that shape the patient-physician dynamic in the context of cardiovascular disease.

## Results

### Study Population Characteristics and Risk Perception

#### Population Characteristics

The study included 943 patients, of whom 315 (33.4%) were women. The mean age was 68.2 years (±14.7). A majority (75.9%) lived with a companion, and a significant portion were retired (62.9%). Most participants were married (61.1%) and had completed primary education (44.1%) [Table 1]. Ischemic heart disease was the leading cause of hospital admission, accounting for 389 patients (41.3%), followed by heart failure in 18.1% [Table 2].

**Table 1.** Patients’ demographics and medical history.

| **Table 1** |  |
| --- | --- |
| **Demographics** | **N = 943** |
| Age, *years* | 68.2 (±14.7) |
| Female sex, *n (%)* | 315 (33.4) |
| Occupation, *n (%)* |  |
| - Retired, *n (%)* | 593 (62.9) |
| - Unemployed, *n (%)* | 60 (6.4) |
| - Actively working, *n (%)* | 229 (24.3) |
| Education |  |
| - Primary education, *n (%)* | 416 (44.1) |
| - Secondary education, *n (%)* | 88 (9.3) |
| - Baccalaureate education, *n (%)* | 121 (12.8) |
| - Vocational training, *n (%)* | 117 (12.4) |
| - University education, *n (%)* | 175 (18.6) |
| - Another education, *n (%)* | 16 (1.7) |
| Civil situation |  |
| - Single, *n (%)* | 124 (13.1) |
| - Married, *n (%)* | 576 (61.1) |
| - Widow, *n (%)* | 146 (15.5) |
| - Divorced, *n (%)* | 62 (6.6) |
| - Another situation, *n (%)* | 21 (2.2) |
| Living with a companion, *n (%)* | 716 (75.9) |
| **Medical history** |  |
| Previous cardiovascular disease, *n (%)* | 439 (46.7) |
| Previous admission due to cardiovascular disease, *n (%)* | 350 (37.1) |
| Family history of cardiovascular disease, *n (%)* | 440 (46.7) |
| Dyslipidaemia, *n (%)* | 363 (38.5) |
| Arterial hypertension, *n (%)* | 488 (51.7) |
| Diabetes mellitus, *n (%)* | 255 (27.0) |
| Smoking (active or former), *n (%)* | 434 (46.0) |
| Overweight or obesity, *n (%)* | 275 (29.2) |

**Table 2.** Patients’ and physicians’ perception and correlation about cause of admission, future cardiovascular risk and explanation of invasive procedures during hospitalization.

| **Table 2** |  |  |  |
| --- | --- | --- | --- |
| **Primary cause of admission** | **Patients' response** | **Physicians' response** | **Kappa Index** |
| Ischaemic heart disease, *n* (%) | 331 (35.1) | 389 (41.3) | 0.72 |
| Heart failure, *n* (%) | 170 (18.0) | 171 (18.1) |  |
| Tachyarrhythmia, *n* (%) | 138 (14.6) | 128 (13.6) |  |
| Bradyarrhythmia, *n* (%) | 36 (3.8) | 61 (6.5) |  |
| Aortic disease, *n* (%) | 9 (1.0) | 4 (0.4) |  |
| Pericardial disease, *n* (%) | 24 (2.5) | 22 (2.3) |  |
| Scheduled procedure, *n* (%) | 99 (10.5) | 84 (8.9) |  |
| Other, *n* (%) | 96 (10.2) | 70 (7.4) |  |
| I don't know, *n* (%) | 22 (2.3) | - |  |
| No opinion/no reply, *n* (%) | 18 (1.9) | 14 (1.5) |  |
| **Future cardiovascular event risk** |  |  | **Weighted Kappa index** |
| Very High, *n* (%) | 90 (9.5) | 192 (20.4) | 0.29 |
| High, *n* (%) | 233 (24.7) | 370 (39.2) |  |
| Moderate, *n* (%) | 114 (12.1) | 205 (21.7) |  |
| Low, *n* (%) | 139 (14.7) | 119 (12.6) |  |
| Very Low, *n* (%) | 73 (7.7) | 47 (5.0) |  |
| I don't know, *n* (%) | 277 (29.4) | 4 (0.4) |  |
| No opinion/no reply, *n* (%) | 17 (1.8) | 6 (0.6) |  |
| **Explanation of procedures (N = 744)** |  |  | **Weighted Kappa index** |
| Strongly agree, *n* (%) | 391 (52.6) | 392 (52.7) | 0.21 |
| Agree, *n* (%) | 181 (24.3) | 302 (40.6) |  |
| I don't know, *n* (%) | 44 (5.9) | 30 (4.0) |  |
| Disagree, *n* (%) | 30 (4.0) | 12 (1.6) |  |
| Strongly disagree, *n* (%) | 70 (9.4) | 2 (0.3) |  |
| No opinion/no reply, *n* (%) | 28 (3.8) | 6 (0.8) |  |

### Patients’ and Physicians’ Risk Perception

#### Understanding of Admission Diagnosis

Patients were asked to identify the primary reason for their hospital admission, and these responses were compared with their treating physicians’ perceptions. Among the 943 patients, 22 (2.3%) were unsure of their admission reason at discharge, and 18 (1.9%) did not respond. The majority of patients (331, 35.1%) identified ischemic heart disease as their reason for admission, followed by heart failure (170, 18.0%). Physicians’ responses closely aligned with those of the patients, resulting in a substantial agreement with a kappa index of 0.72 [Table 2, Central Illustration].

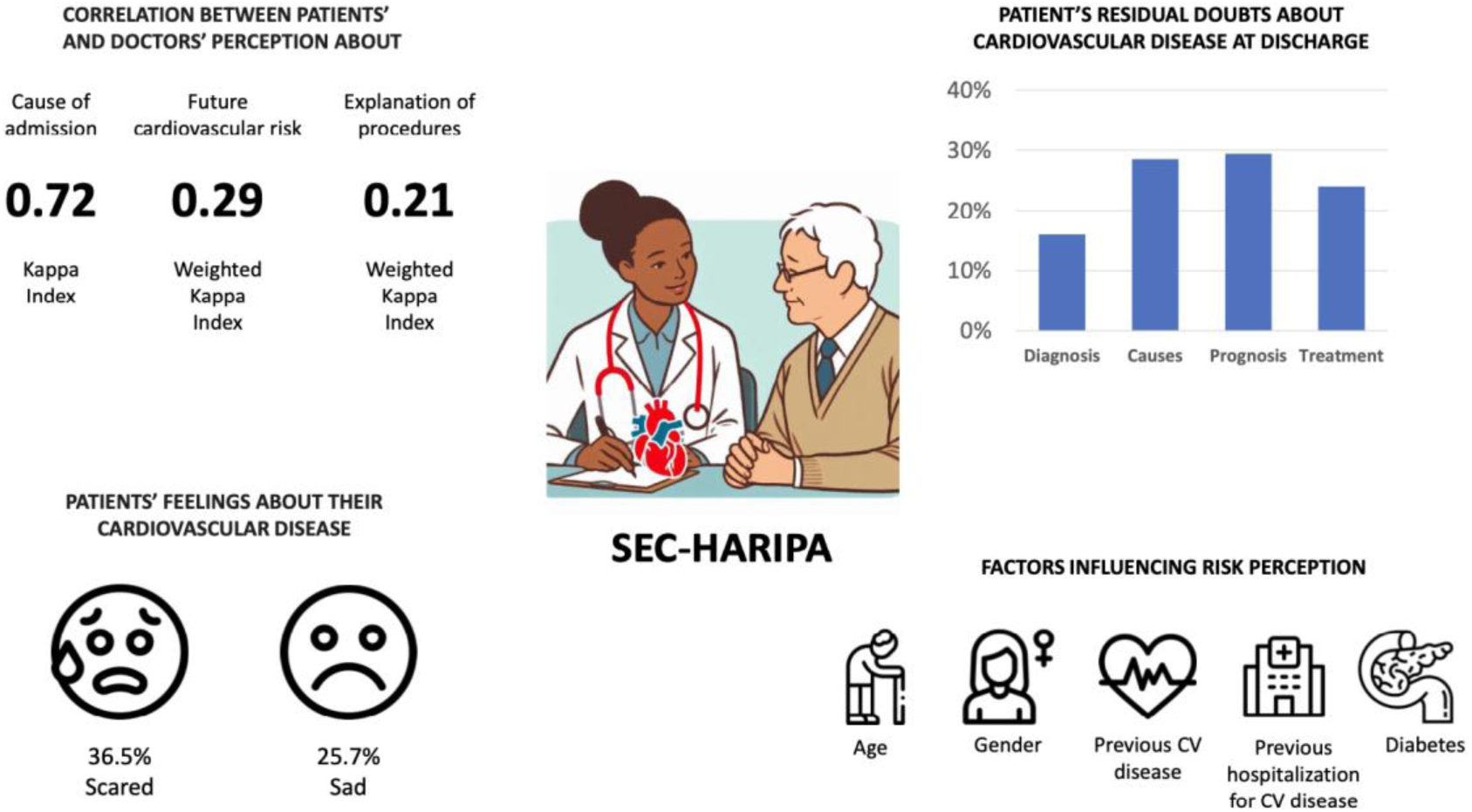
CENTRAL ILLUSTRATION

#### Perceived Risks of Future Events

Patients’ perceptions of their future cardiovascular event risk were assessed using a visual scale. A significant number of patients (277, 29.4%) reported being unaware of their future cardiovascular risk, with only 17 (1.8%) leaving this question unanswered. Among those who provided an assessment, most perceived their risk as high (233, 24.7%) or very high (90, 9.5%). However, physicians often rated the patients’ risk higher, with a significant proportion indicating high (192, 20.4%) or very high (370, 39.2%) risk. The correlation between patients’ and physicians’ risk perceptions was low, with a weighted kappa index of 0.29, indicating that patients generally underestimated their risk compared to their physicians’ assessments [Table 2, Central Illustration].

#### Communication About Complications of Interventions

When asked about the information they received regarding potential complications associated with their interventions, there was a notable discrepancy between patients and physicians. A majority of patients (572, 76.9%) agreed that they had received this information, while a small but significant portion (100, 13.4%) disagreed, and 72 (9.7%) were uncertain or did not respond. Conversely, physicians overwhelmingly reported that they had communicated this information, with 669 (93.4%) stating they “strongly agree” or “agree” that they had explained the risks. However, the correlation between the patients’ and physicians’ responses was low, with a weighted kappa index of 0.21, highlighting a significant gap in the perceived communication of risk information [Table 2, Central Illustration].

#### Information About Complication Risks from Invasive Procedures

The questionnaire assessed patients’ perceptions of the information they received regarding potential complications from invasive procedures during their hospital stay. Of the 943 participants, 744 (78.9%) underwent an intervention (a total of 888 invasive procedures were performed) [Table 3]. Among these, 572 (76.9%) “strongly agreed” or “agreed” that they received adequate information about risks. However, 100 patients (13.4%) “disagreed” or “strongly disagreed,” and 72 (9.7%) were uncertain or did not respond [Table 3]. Notably, among the 199 patients who experienced adverse events related to their intervention, a majority (138, 69.3%) indicated they had not been informed about the possibility of such events.

**Table 3.** Invasive procedures and new medication during hospitalization and explanation of possible adverse events.

| **Table 3** |  |
| --- | --- |
| **Invasive procedures performed** | **N = 888** |
| Cardiac catheterization (with or without percutaneous coronary intervention), *n* (%) | 501 (47.9) |
| Transcatheter aortic valve implantation, *n* (%) | 48 (5.0) |
| Arrhythmia ablation, *n* (%) | 58 (6.0) |
| Pacemaker or defibrillator implantation, *n* (%) | 115 (11.9) |
| Electric cardioversion, *n* (%) | 43 (4.4) |
| Cardiac surgery (valvular replacement, coronary bypass graft surgery...), *n* (%) | 13 (1.3) |
| Ventricular assist device implantation, *n* (%) | 4 (0.4) |
| Others, *n* (%) | 106 (11.0) |
| **In case of adverse events, explanation before the procedure** | **N = 199** |
| Yes, *n* (%) | 61 (30.7) |
| No, *n* (%) | 138 (69.3) |
| **New medication prescribed during hospitalization** | **N = 943** |
| Yes, *n* (%) | 643 (68.2) |
| No, *n* (%) | 276 (29.3) |
| No opinion/no reply, *n* (%) | 24 (2.5) |
| **Explanation of possible adverse reactions** | **N = 643** |
| Strongly agree, *n* (%) | 228 (35.5) |
| Agree, *n* (%) | 133 (20.7) |
| I don't know, *n* (%) | 82 (12.8) |
| Disagree, *n* (%) | 67 (10.4) |
| Strongly disagree, *n* (%) | 117 (18.2) |
| No opinion/no reply, *n* (%) | 16 (2.5) |
| **In case of adverse reaction, explanation before administrate the medication** | **N = 179** |
| Yes, *n* (%) | 91 (50.8) |
| No, *n* (%) | 88 (49.2) |

#### Information About Medication

Among the 643 patients (68.2%) who reported receiving new medications during their admission, the majority either “agreed” (133, 20.7%) or “strongly agreed” (228, 35.5%) that they received adequate information about potential adverse events. However, 16 patients (2.5%) did not respond [Table 3]. Notably, among the 179 patients who experienced adverse events related to their medication, nearly half (88, 49.2%) indicated they were not informed about the possibility of such events [Table 3].

#### Residual Doubts Related to Cardiovascular Disease

The questionnaire explored patients’ residual doubts about their cardiovascular disease. Specifically, 151 patients (16%) expressed doubts about the disease for which they were admitted, 269 (28.5%) had uncertainties about its causes, 277 (29.4%) were unsure about the future prognosis, and 226 (24.0%) had lingering questions about their treatment [Table 4, Central Illustration].

**Table 4.** Patients’ doubts at discharge and emotions about their cardiovascular disease.

| **Table 4** |  |
| --- | --- |
| **Residual doubts about own disease at discharge** | **N = 943** |
| Diagnosis, n (%) | 151 (16.0) |
| Causes, n (%) | 269 (28.5) |
| Prognosis, n (%) | 277 (29.4) |
| Treatment, n (%) | 226 (24.0) |
| Risks from treatment, n (%) | 258 (27.4) |
| **Patients' feelings about their cardiovascular risk** |  |
| Sad, n (%) | 242 (25.7) |
| Eh/indifferent, n (%) | 74 (7.8) |
| Confident, n (%) | 120 (12.7) |
| Scared, n (%) | 344 (36.5) |
| Confused, n (%) | 91 (9.7) |
| Mad, n (%) | 54 (5.7) |
| No opinion/no reply, n (%) | 18 (1.9) |
| **Patients' feelings about their ability to modify cardiovascular risk** |  |
| Sad, n (%) | 70 (7.4) |
| Eh/indifferent, n (%) | 70 (7.4) |
| Confident, n (%) | 654 (69.4) |
| Scared, n (%) | 51 (5.4) |
| Confused, n (%) | 64 (6.8) |
| Mad, n (%) | 24 (2.5) |
| No opinion/no reply, n (%) | 10 (1.1) |

### Patients’ Feelings

The survey assessed patients’ emotions regarding their cardiovascular risk. The most frequently reported emotion was “scared,” noted by 344 patients (36.5%), followed by “sad” in 242 patients (25.7%). When considering their ability to take preventive actions against heart problems, a majority felt “confident” (654, 69.4%), with smaller proportions feeling “sad” (70, 7.4%) or “indifferent” (70, 7.4%) [Table 4, Central Illustration].

### Factors Influencing Risk Perception

Several factors significantly influenced patients’ perceptions of their cardiovascular risk. Among the participants, 439 (46.6%) had a history of cardiovascular disease (CVD), and 350 (37.1%) had been previously hospitalized due to a cardiovascular event [Table 5]. The presence of a prior cardiovascular condition was independently associated with a higher self-perception of risk (OR 4.71, 95% CI 3.03-7.33, p<0.001), as was having a previous cardiovascular hospitalization (OR 3.00, 95% CI 1.63-5.53, p<0.001). Patients with a history of CVD or previous cardiovascular hospitalization had a risk perception more closely aligned with their treating physicians, as indicated by a higher kappa index (CVD history: KI 0.36; No history: KI 0.15; Prior hospitalization: KI 0.27; No prior hospitalization: KI 0.24).

**Table 5.**
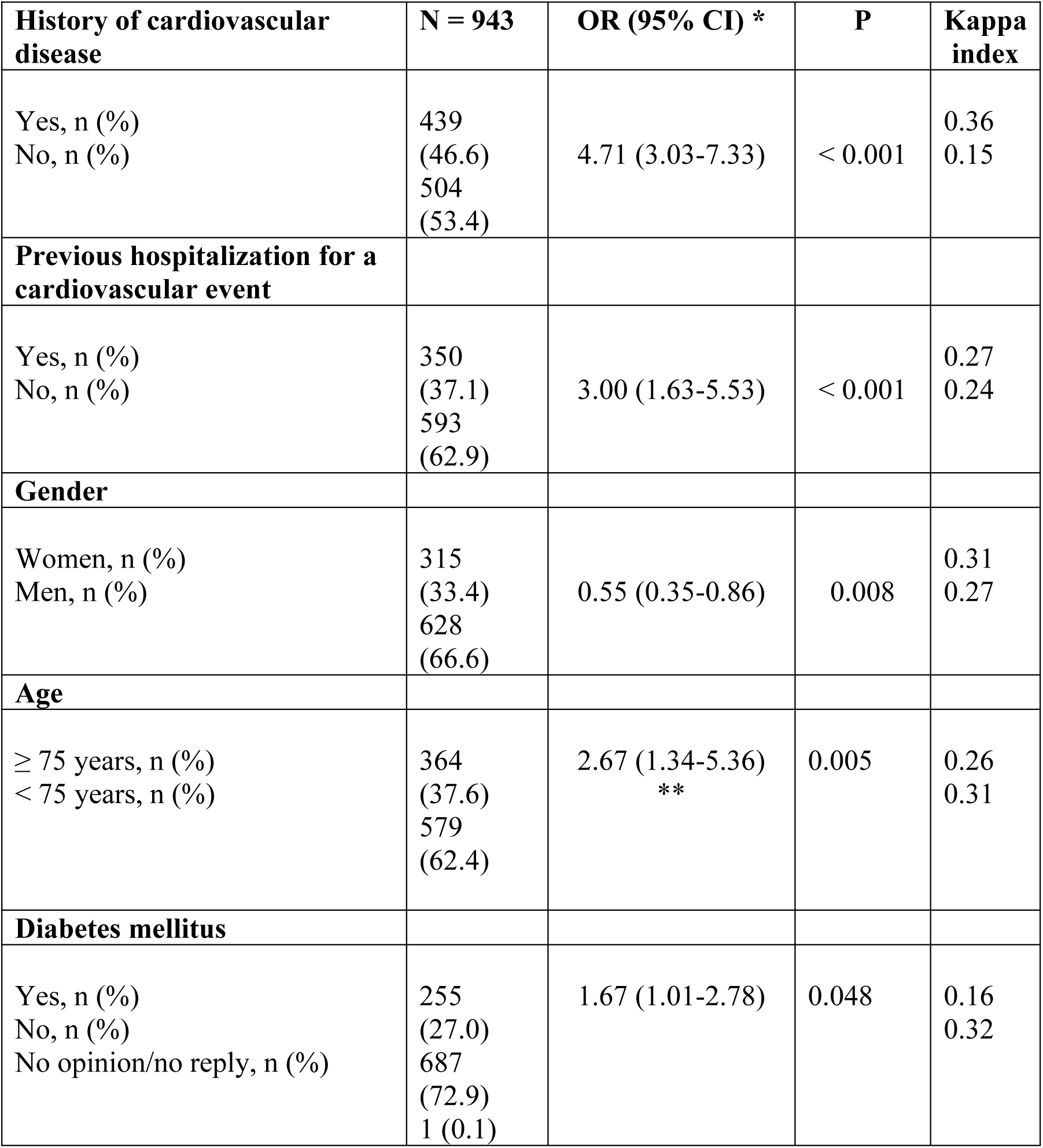
Factors influencing risk perception. *Very high/high-risk and low/very low-risk of patients’ risk of having another cardiovascular event or heart-related procedure/device (**Very high-risk and very low-risk)

| **Table 5** |  |  |  |  |
| --- | --- | --- | --- | --- |
| **History of cardiovascular disease** | **N = 943** | **OR (95% CI) \*** | **P** | **Kappa index** |
| Yes, n (%)<br>No, n (%) | 439<br>(46.6)<br>504<br>(53.4) | 4.71 (3.03-7.33) | < 0.001 | 0.36<br>0.15 |
| **Previous hospitalization for a cardiovascular event** |  |  |  |  |
| Yes, n (%)<br>No, n (%) | 350<br>(37.1)<br>593<br>(62.9) | 3.00 (1.63-5.53) | < 0.001 | 0.27<br>0.24 |
| **Gender** |  |  |  |  |
| Women, n (%)<br>Men, n (%) | 315<br>(33.4)<br>628<br>(66.6) | 0.55 (0.35-0.86) | 0.008 | 0.31<br>0.27 |
| **Age** |  |  |  |  |
| ≥ 75 years, n (%)<br>< 75 years, n (%) | 364<br>(37.6)<br>579<br>(62.4) | 2.67 (1.34-5.36)<br>** | 0.005 | 0.26<br>0.31 |
| **Diabetes mellitus** |  |  |  |  |
| Yes, n (%)<br>No, n (%)<br>No opinion/no reply, n (%) | 255<br>(27.0)<br>687<br>(72.9)<br>1 (0.1) | 1.67 (1.01-2.78) | 0.048 | 0.16<br>0.32 |

### Emotional Responses

Emotional responses also played a role in shaping risk perception. Patients without prior cardiovascular hospitalizations predominantly reported feeling “scared” (41.1%), followed by “sadness” (23.6%). In contrast, those with prior hospitalizations had a more balanced distribution of “fear” (30.5%) and “sadness” (30.2%) (p<0.001). Patients with doubts about the cause of their admission or their treatment were more likely to report feeling scared, while those without such doubts more frequently reported feeling sad. Additionally, both being aged 75 or older and having a previous cardiovascular hospitalization were independent predictors and protective factors against experiencing fear and sadness. Undergoing catheterization during the hospital stay was also identified as an independent predictor of fear.

### Gender Differences

Gender also influenced risk perception. Among the study population, 315 (33.4%) were women [Table 5]. Men were more likely to perceive their adjusted risk of a future cardiovascular event as lower (high/very high vs. low/very low) compared to women (OR 0.55, 95% CI 0.35-0.86, p=0.008). The correlation between men’s risk perceptions and their physicians’ and women’s risk perceptions and their physicians’ was lower (KI 0.27 and KI 0.31 respectively).

### Age-Related Differences

Age was another significant factor. Among the participants, 364 (37.6%) were 75 years or older [Table 5]. Compared to younger patients, those 75 and older had undergone more valve implantations and electric device procedures but fewer catheterizations and arrhythmia ablations (3.4% vs. 7.7%, p=0.006). Older patients were also more likely to perceive a greater risk of a recurrent cardiovascular event, especially in extreme response categories (Very High and Very Low Risk) (OR 2.67, 95% CI 1.34-5.36, p=0.005). Younger patients (under 75) more frequently reported feeling “scared” (247; 43.3%), followed by “sad” (140; 24.6%), whereas older patients more commonly reported feeling “sad” (102; 28.9%), followed by “scared” (97; 27.5%).

### Diabetes Mellitus

The presence of diabetes mellitus (DM) was independently associated with a higher self-perception of cardiovascular recurrence risk compared to those without DM (OR 1.67, 95% CI 1.01-2.78, p=0.048) [Table 5].

## Discussion

This study offers a comprehensive exploration of the complex dynamics of risk perception and communication between physicians and patients in a real-world cardiovascular setting. Our findings underscore significant gaps in communication, particularly in how patients perceive their cardiovascular risk and the information they receive about procedural complications.

### Study Population Characteristics and Risk Perception

The patient population in this study is a microcosm of the broader cardiovascular landscape, with ischemic heart disease, heart failure, and arrhythmias standing out as the predominant causes of admission^19^. The high rate of invasive procedures (78.9%) observed among these patients mirrors the intensive nature of contemporary cardiovascular care, where such interventions are routine. Yet, our findings reveal a critical gap in communication: more than one in five patients (21.1%) either disagreed with having received sufficient information about these procedures, were unsure, or did not respond. Furthermore, among those who experienced adverse events, a striking 69.3% reported that they had not been informed of potential complications beforehand.

These results are particularly concerning given the established importance of effective RC in patient care. Previous studies, such as those by Fagerlin et al.^20^, have shown that patients often struggle with understanding medical risks, leading to significant discrepancies between what physicians believe they have communicated and what patients actually comprehend. This disconnect can have serious implications, not only for patient satisfaction but also for adherence to treatment and overall outcomes.

The discharge process emerged as another critical area for improvement. Effective discharge planning, including thorough patient education, is associated with better long-term outcomes and reduced risk of recurrent events ^21,22^. However, our data indicate that many patients leave the hospital with lingering uncertainties about their prognosis (29.4%), treatment plans (24.0%), and the causes of their condition (28.5%). These findings highlight a significant opportunity to enhance discharge education and ensure that patients are adequately prepared to manage their health post-hospitalization. The emotional toll of cardiovascular disease is also evident, with over a third of patients (36.5%) expressing fear of future events and a quarter (25.7%) reporting feelings of sadness at discharge. These emotions, if not properly addressed, could undermine patient recovery and long-term adherence to treatment plans.

### Patients’ Risk Perception and Physicians’ Risk Perception

One of the most compelling aspects of this study is the dual-perspective approach, allowing us to directly compare patient and physician perceptions of risk communication. The alignment between patients and physicians regarding the primary reason for admission was relatively strong, with a kappa index of 0.72. This suggests that, at least in terms of diagnosis, there is a shared understanding between the two groups. This level of agreement is encouraging and consistent with previous research indicating that clear communication about the reason for hospitalization is generally well-conveyed.

However, the study uncovers significant discrepancies in other areas. Despite 93.4% of physicians believing they had effectively communicated procedural risks, only 76.9% of patients agreed, resulting in a low weighted kappa index of 0.21 (Table 3). This mismatch points to a deeper issue in how risk information is conveyed and received, echoing findings from studies like those by Mazor et al ^23^, which suggest that even when physicians feel they have communicated effectively, patients may still be left with misunderstandings or doubts.

The discordance extends to future cardiovascular risk assessments, where the correlation between patient and physician perspectives was similarly low (weighted kappa index of 0.29). This discrepancy is not unique to our study; a meta-analysis by Bakhit et al ^1^ found that patients often struggle to accurately assess their cardiovascular risk, even when provided with detailed risk information. Patients frequently underestimated their risk compared to physician assessments, a trend that aligns with the concept of “optimistic bias,” where individuals perceive their personal risk as lower than it objectively is ^24^. This underestimation is particularly problematic in cardiovascular care, where accurate risk perception is crucial for motivating lifestyle changes and adherence to preventive therapies. As highlighted by Navar et al^25^, effective communication in cardiovascular disease prevention is not just about what is said but also how it is said. Their model emphasizes the importance of assessing patient priorities, clearly communicating risks and benefits, and engaging in shared decision-making to align patient and physician perspectives effectively.

### Factors Influencing Risk Perception

In exploring factors that influence risk perception, our analysis identified several key determinants. Patients with a history of cardiovascular disease or prior hospitalizations were more likely to have a higher perception of future risk, and their perceptions more closely aligned with those of their physicians. This finding is consistent with prior research indicating that personal experience with a disease heightens awareness and concern about future risks^26^. Similarly, patients with diabetes mellitus, a condition that exacerbates cardiovascular risk, also demonstrated a heightened risk perception.

Gender and age were also significant factors in risk perception. Women and older adults (aged 75 and above) showed a higher perception of risk, with better alignment with physician assessments compared to men and younger patients. This may reflect broader gender and age-related differences in health behaviors and risk communication, as women are often more proactive in health-related discussions and older adults may perceive themselves as more vulnerable ^27^.

These findings underscore the need for personalized communication strategies that account for individual patient characteristics, as emphasized in the the meta-analysis by Bakhit et al ^1^. By tailoring risk communication to the specific needs and backgrounds of patients, we can improve their understanding, reduce emotional distress, and enhance their ability to make informed decisions about their health. This approach is essential for achieving better alignment between patient perceptions and clinical realities, ultimately leading to improved outcomes in cardiovascular care.

This study offers valuable insights into the dynamics of RC between physicians and patients in the context of cardiovascular disease; however, several limitations should be acknowledged. The cross-sectional nature of the study limits the ability to infer causality between the observed discrepancies in risk perception and outcomes. Additionally, the study focuses on a single hospital admission, which may not capture the full spectrum of RC that occurs across multiple interactions over time. Patients’ perceptions of risk and their understanding may change with subsequent visits, follow-ups, or after additional education, which this study does not assess. While the study provides a dual perspective by comparing patient and physician perceptions, we did not explore the underlying reasons for the discrepancies observed. Furthermore, the study population includes a significant number of elderly patients, but it does not thoroughly account for the impact of cognitive impairments, emotional distress, or geriatric syndromes on risk perception and communication. These factors can substantially influence how information is processed and understood, particularly in older adults. Although the questionnaire was carefully translated and adapted for the Spanish-speaking population, the study may not fully address the cultural nuances that influence risk perception and communication. Differences in health literacy and cultural attitudes towards illness and risk could affect the generalizability of the findings to other populations or settings.

Finally, the study relied on patients’ self-reported perceptions and recollections of the information provided by physicians, which may introduce recall bias, especially given that patients were surveyed close to discharge when their emotional and cognitive states might have been affected by their recent medical experience.

## Conclusions

The SEC-HARIPA study highlights critical gaps in risk communication between patients and physicians during cardiovascular care. First, many patients underestimated their cardiovascular risk and left the hospital with unresolved doubts, indicating a need for clearer communication. Second, there was a significant misalignment between patients’ and physicians’ perceptions of risk, especially regarding future events and procedural complications. Third, factors such as prior cardiovascular history, gender, and age influenced how patients perceived their risk, emphasizing the importance of personalized communication strategies.

Overall, improving the clarity and personalization of RC can enhance patient understanding, align perceptions with clinical realities, and ultimately lead to better health outcomes in cardiovascular care. Future efforts should focus on tailored communication approaches, particularly for vulnerable groups like the elderly.

## Data Availability

The data used in this study are available upon reasonable request, subject to [privacy restrictions, confidentiality agreements, etc.]. Interested parties can contact the corresponding author at to obtain access to the relevant data.

## Abbreviations list

DM: diabetes mellitus
RC: Risk Communication

## Acknowledgments

Thank the Spanish Society of Cardiology for promoting the conduct of this study. Additionally, thank all the research teams from the 28 hospitals that made it possible to collect this data and obtain the subsequent results.

## Declaration of interest statement

All authors have reported that they have no relationships relevant to the contents of this paper to disclose.

## Notes

### Competing Interest Statement

The authors have declared no competing interest.

### Funding Statement

- The authors declare that they did not receive any payments or services from third parties for any part of the submitted work, including study design, manuscript preparation, or statistical analysis. - his study did not receive external funding.

### Author Declarations

The research protocol received approval from the Ethics Committee of the Central University Hospital of the Defense "Gómez Ulla", as well as from the Ethics Committees of the participating hospitals, and adhered to the Helsinki Protocol.

